# Nomogram for the Prediction of Shunt-Dependent Hydrocephalus in Patients with Aneurysmal Subarachnoid Hemorrhage: A Single-institute Experience

**DOI:** 10.1101/2022.12.31.22283967

**Authors:** Chia-Ryo Wu, Jin-Shuen Chen, Yao-Shen Chen, Chun-Hao Yin, Wei-Chuan Liao, Yu-Lun Wu, Yu-Hone Hsu

## Abstract

**BACKGROUND:** This study is focused to identify the risk factors of shunt-dependent hydrocephalus (SDHC) after aneurysmal subarachnoid hemorrhage (aSAH) and develop a model to predict its incidence.

**METHODS:** Medical records of 118 consecutive patients with aSAH treated in our institution from January 2013 to October 2021 were reviewed retrospectively, 109 of them were enrolled in this cohort, the following data were analyzed based on presence or absence of SDHC: age, gender, body mass index, Fisher grade, Hunt and Hess scale (HHS), aneurysm location, treatment modality, new neurological deficits after aneurysm treatment, estimated glomerular filtration rate (eGFR), neutrophil-lymphocyte ratio (NLR), platelet-lymphocyte ratio (PLR), and platelet-neutrophil ratio (PNR). We conducted univariate and multivariate logistic regression analyses to illustrate a nomogram for predicting SDHC risk.

**RESULTS:** The stepwise logistic regression analysis with backward selection revealed three independent predictive factors of SDHC: age ≥65 (odds ratio, 3.94; 95% CI, 1.4–11.00; *p* = 0.009), treatment modality (odds ratio, 4.36; 95% CI, 1.81–10.53; *p* = 0.001), and HHS ≥3 (odds ratio, 3.59; 95% CI, 1.50–8.61; *p* = 0.004). A nomogram for SDHC risk prediction was developed based on the weight of these 3 factors.

**CONCLUSIONS:** Age, treatment modality (clipping vs coiling), and HHS are predictive for SDHC after aSAH. Endovascular embolization of aneurysm plays an important role in reducing risk of SDHC after aSAH.

## INTRODUCTION

Shunt-dependent hydrocephalus (SDHC) is one of the clinical problems that cause extended hospital and intensive care unit stays in patients with aneurysmal subarachnoid hemorrhage (aSAH).^1^ Symptomatic hydrocephalus is usually recognized as an important cause of diminished consciousness and manifestation of intracranial hypertension, which usually associates with poor outcomes. Ventriculoperitoneal (VP) shunt is the most usual treatment in patients with aSAH needing cerebrospinal fluid (CSF) diversion. Owing to the unignorable complications in VP shunt placement, the clinical treatment to reduce SDHC risk gains increasing concerns in the current aneurysm treatment.

The risk factors of SDHC after aSAH have been widely reported, including old age^4,6,8,14,17,19^, female gender^8^, BMI^10^, Hunt and Hess scale (HHS)^4,6,8,9,13,15,17,19,28^, Fisher’s grade^4,6,7,8,9,15,17,19^ and aneurysm location^6,8,28^, estimated glomerular filtration rate (eGFR)^5^, new neurological deficit secondary to infarction, vasospasm and rebleeding^6,8,24,28^, treatment modality (surgical clipping vs. endovascular coiling)^2,8,14,15,16,17,18,19,20,21,22,23,24,25^. Besides, neutrophil-lymphocyte ratio (NLR), platelet-lymphocyte ratio (PLR), and platelet-neutrophil ratio (PNR) are associated with clinical outcomes of aSAH^26,27^. Despite the risk factors reported, only a few studies have developed a prediction tool for risk assessment of SDHC in patients with aSAH after aneurysm treatment. Thus, this study aimed to create a predictive model for SDHC in patients with aSAH, which may help early recognize patients needing VP shunt surgery and optimize treatment result.

## MATERIALS AND METHODS

This study was approved by institutional review board of Kaohsiung Veterans General Hospital Medical record of 118 consecutive patient who underwent aneurysm obliteration treatment following aSAH in our institution from January 2013 to October 2021 were reviewed retrospectively, all patients had brain CTA with or without cerebral catheter angiography study to confirm the diagnosis of aSAH, 8 patients with in-hospital mortality after aneurysm treatment and 1 patient with incomplete laboratory data were excluded, finally 109 patients were enrolled in this cohort.

The following data were analyzed based on presence or absence of SDHC: age, gender, BMI, aneurysm treatment modality (clipping or coiling), new neurological deficits after aneurysm treatment due to infarction, rebleeding or delayed cerebral ischemia, on admission laboratory data including eGFR, NLR, PLR and PNR, location of aneurysm (anterior or posterior circulation), Fisher grade and HHS before aneurysm treatment.

The decision of aneurysm treatment modality was based on the consensus between neurosurgeons and neurointerventionalists for each patient, treatment was carried out within 72 hours after bleeding to minimize risk of rebleeding. Every patient had external ventricular drainage (EVD) surgery on admission or during the surgical clipping procedures, or immediately after endovascular coiling.

All the patients received EVD challenge between 1–4 weeks after aSAH, the EVD challenge protocol was EVD closure with continuous intracranial pressure (ICP) monitoring for 72 h. If there was any ICP episode elevation over 20 mmHg for more than 5 min, clinical deterioration (headache, nausea, vomiting, decreased mentality, or Glasgow Coma Scale), or hydrocephalus status worsening in the CT scan performed at the end of challenge, the protocol would be considered failed. Patients with failed EVD challenge and those who developed hydrocephalus after successful EVD removal underwent VP shunt surgery.

### Statistical analysis and predictive nomogram for SDHC risk

We analyzed categorical variables of SDHC, with the chi-square test and Fisher’s exact test for SDHC. Continuous variables expressed as means ± standard deviations were compared using an independent t test. All variables with *p* < 0.1 in the univariate analysis were subsequently included in the multivariate logistic regression analyses. The results of the comparison are expressed using *p* values and *p* < 0.05 was considered significant propensity. We also constructed a nomogram to estimate SDHC risk plotted to determine the numerical predictors of SDHC based on significant variables selected from the multivariate logistic regression model.

The cutoff value for continuous variables in predicting SDHC was determined if they received operating characteristic curves (ROC), a cutoff value set with a highest area under the curve (AUC). We ranked the estimated effects of each variable with estimated beta coefficients and plotted calibration curves to assess the nomogram calibration, along with the Hosmer–Lemeshow test and an area under the receiver operating characteristic curve (AUC) for predictive performance. All statistical analyses were conducted with the SAS software package, version 9.4 (SAS Statistical Institute, Cary, NC, USA) and SPSS for Windows version 20 (SPSS Inc, Chicago, IL, USA).

## RESULTS

### Baseline characteristics of patients and SDHC predictors

A total of 118 consecutive patients underwent surgical clipping or endovascular coiling for ruptured aneurysm treatment. Eight patients who died within 7 days and one patient who lacked complete data were excluded from this study. Finally, 109 patients were enrolled in this study. The demographic data is shown in **Table 1**. The median patient age was 56.8 years, gender distribution was male 29% (32 patients), female 71% (77 patients). In 53 (49%) patients, ruptured aneurysms were repaired by coiling, the other 56 (51%) patients were repaired by clipping. 28 of 109 patients (26%) developed new neurological deficits after aneurysm treatment. The responsible aneurysms were located in the anterior circulation in 81 patients (74%) and in the posterior circulation in 26 patients (26%), the Fisher grade was 2 in 5 patients (4.5%), 3 in 39 patients (35.8%), and 4 in 65 patients (59.6%). The HHS distribution 1–5 was 10 patients (9%), 47 patients (43%), 12 patients (11%), 29 patients (27%), and 11 patients (10%), respectively.

**Table 1.** Baseline Characteristics of Aneurysmal Subarachnoid Hemorrhage Patients. (*n*=109)

|  |  |  |  |  |
| --- | --- | --- | --- | --- |
| Variable | Total<br>n=109 (%) | V-P shunt<br>n=50 (%) | No V-P shunt<br>n=59 (%) | p-value |
| **Age (years) (mean ± SD)** | 56.8±13.0 | 60.3±14.5 | 53.9±11.0 | 0.009 |
| **Gender** |  |  |  | 0.892 |
| Male | 32 (29%) | 15 (30%) | 17 (29%) |  |
| Female | 77 (71%) | 35 (70%) | 42 (71%) |  |
| **BMI (mean ± SD)** | 23.1±4.1 | 23.0±2.9 | 23.2±4.9 | 0.760 |
| **Treatment** |  |  |  | 0.001 |
| Coiling | 53 (49%) | 16 (32%) | 37 (63%) |  |
| Clipping | 56 (51%) | 34 (68%) | 22 (37%) |  |
| **New neurological deficit** |  |  |  | 0.067 |
| Yes | 28 (26%) | 17 (34%) | 11 (19%) |  |
| No | 81 (74%) | 33 (66%) | 48 (81%) |  |
| **eGFR (ml/min/1.73%) (mean ±SD)** | 88.8±23.5 | 86.5±21.8 | 90.7±24.9 | 0.356 |
| **NLR (mean ± SD)** | 9.7±11.8 | 10.0±9.8 | 9.4±13.3 | 0.799 |
| **PLR (mean ± SD)** | 204.5±189.1 | 202.1±158.1 | 206.5±213.1 | 0.903 |
| **PNR (mean ± SD)** | 29.3±16.6 | 26.4±15.8 | 31.8±16.9 | 0.090 |
| **Location** |  |  |  | 0.611 |
| Anterior circulation | 81 (74%) | 36 (72%) | 45 (76%) |  |
| Posterior circulation | 28 (26%) | 14 (28%) | 14 (24%) |  |
| **Fisher grade** |  |  |  | 0.102 |
| 2 | 5 (5%) | 1 (2%) | 4 (7%) |  |
| 3 | 39 (36%) | 14 (28%) | 25 (42%) |  |
| 4 | 65 (60%) | 35 (70%) | 30 (51%) |  |
| **Hunt and Hess scale** |  |  |  | 0.001 |
| 1 | 10 (9%) | 4 (8%) | 6 (10%) |  |
| 2 | 47 (43%) | 15 (30%) | 32 (54%) |  |
| 3 | 12 (11%) | 3 (6%) | 9 (15%) |  |
| 4 | 29 (27%) | 19 (38%) | 10 (17%) |  |
| 5 | 11 (10%) | 9 (18%) | 2 (3%) |  |
| Abbreviation: SDHC, shunt-dependent hydrocephalus; OR, odd ratio; SD, standard deviation; BMI, Body Mass Index; eGFR, estimated glomerular filtration rate; AH, Acute hydrocephalus; EVD, External ventricular drain; NLR, Neutrophil-lymphocyte ratio; PLR, Platelet-lymphocyte ratio; PNR, Platelet-neutrophil ratio |  |  |  |  |

In the univariate analysis **(Table 2)**, the following three variables were significantly different between the V-P shunt and no V-P shunt groups: age ≥65 years (odds ratio [OR], 3.13; 95% confidence interval [CI], 1.25–7.80; *p =* 0.015), treatment (clipping versus coiling) (OR, 3.57; 95% CI, 1.61–7.91; *p =* 0.002), and HHS ≥3 (OR, 2.95; 95% CI, 1.35–6.45; *p =* 0.007). Regarding the above three risk factors, significance and clear cutoff values were presented with an area under the curve value of 0.753. The stepwise logistic regression analysis showed that age ≥65 years (OR, 3.94; 95% CI, 1.41–11.00; *p =* 0.009), treatment (clipping versus coiling) (OR, 4.36; 95% CI, 1.81–10.53; *p* = 0.001), and HHS ≥3 (OR, 3.59; 95% CI, 1.50–8.61; *p =* 0.004) contributed to SDHC development **(Table 3)**. The AUC (c-statistic) for this model in predicting SDHC of the nomogram had a value of 0.753; 95% CI, 0.66–0.84.

**Table 2.** Univariate Logistic Regression Analysis of Predictor Factor for Shunt-Dependent Hydrocephalus After Aneurysmal Subarachnoid Hemorrhage (*n*=109)

|  |  |  |  |
| --- | --- | --- | --- |
| Variables | Beta | OR (95% CI) | *p* value |
| **Age (years) $\geq 65$** | 1.139 | 3.13 (1.25–7.80) | 0.015 |
| **Gender, male** | 0.057 | 1.06 (0.46–2.42) | 0.892 |
| **BMI** | -0.015 | 0.99 (0.90–1.08) | 0.758 |
| **Treatment** |  |  |  |
| Clipping versus coiling | 1.274 | 3.57 (1.61–7.91) | 0.002 |
| **New neurological deficit** | 0.810 | 2.25 (0.93–5.41) | 0.071 |
| **eGFR (ml/min/1.73%) $< 60$** | 0.629 | 1.88 (0.50–7.06) | 0.353 |
| **NLR $\geq 6.8$** | 0.330 | 1.39 (0.65–2.96) | 0.392 |
| **PLR $\geq 160$** | 0.262 | 1.30 (0.61–2.77) | 0.496 |
| **PNR $\geq 25$** | -0.575 | 0.56 (0.26–1.21) | 0.140 |
| **Location** |  |  |  |
| Posterior circulation versus Anterior circulation | 0.223 | 1.25 (0.53–2.96) | 0.611 |
| **Fisher grade** |  |  |  |
| $\geq 3$ | 0.806 | 2.24 (0.23–22.05) | 0.489 |
| $\geq 4$ | 1.540 | 4.67 (0.49–44.05) | 0.179 |
| **Hunt and Hess scale $\geq 3$** | 1.083 | 2.95 (1.35–6.45) | 0.007 |
| Abbreviation: SDHC, shunt-dependent hydrocephalus; OR, odd ratio; SD, standard deviation; BMI, Body Mass Index; eGFR, estimated glomerular filtration rate; AH, Acute hydrocephalus; EVD, External ventricular drain; NLR, Neutrophil-lymphocyte ratio; PLR, Platelet-lymphocyte ratio; PNR, Platelet-neutrophil ratio |  |  |  |

**Table 3.** Stepwise Logistic Regression Analysis for Shunt-Dependent Hydrocephalus After Aneurysmal Subarachnoid Hemorrhage (*n*=109)

|  |  |  |  |
| --- | --- | --- | --- |
| Variables | Beta | OR (95% CI) | *p* value |
| **Age (years) $\geq 65$** | 1.371 | 3.94 (1.41–11.00) | 0.009 |
| **Treatment** |  |  |  |
| Clipping versus coiling | 1.472 | 4.36 (1.81–10.53) | 0.001 |
| **HHS $\geq 3$** | 1.279 | 3.59 (1.50–8.61) | 0.004 |
| **c-statistic** | | 0.753 (0.66–0.84) | $<0.001$ |
| **HL test** |  | Chi-square test=0.995 | 0.963 |
| Abbreviation: SDHC, shunt-dependent hydrocephalus; SD, standard deviation; OR, odds ratio; CI, confidence interval; HL, Hosmer–Lemeshow; HHS, Hunt and Hess scale |  |  |  |

### Prognostic nomogram for predictors of SDHC development

According to the results of the multivariate logistic regression, the nomogram for the development of predictive scoring systems is feasible. To illustrate the prognostic nomogram, stepwise logistic regression analysis defined each significant variable (age, treatment, and HHS) as 1 point on the point scale. A total score was depicted after the definition of each point from each factor. After locating and summating each score on the total point scale, one straight line could be drawn for the estimation of SDHC probability. The nomogram is depicted in **Figure 1**, with the Hosmer–Lemeshow test values (Chi-square test = 0.995; *p* = 0.963).

**Figure 1.**
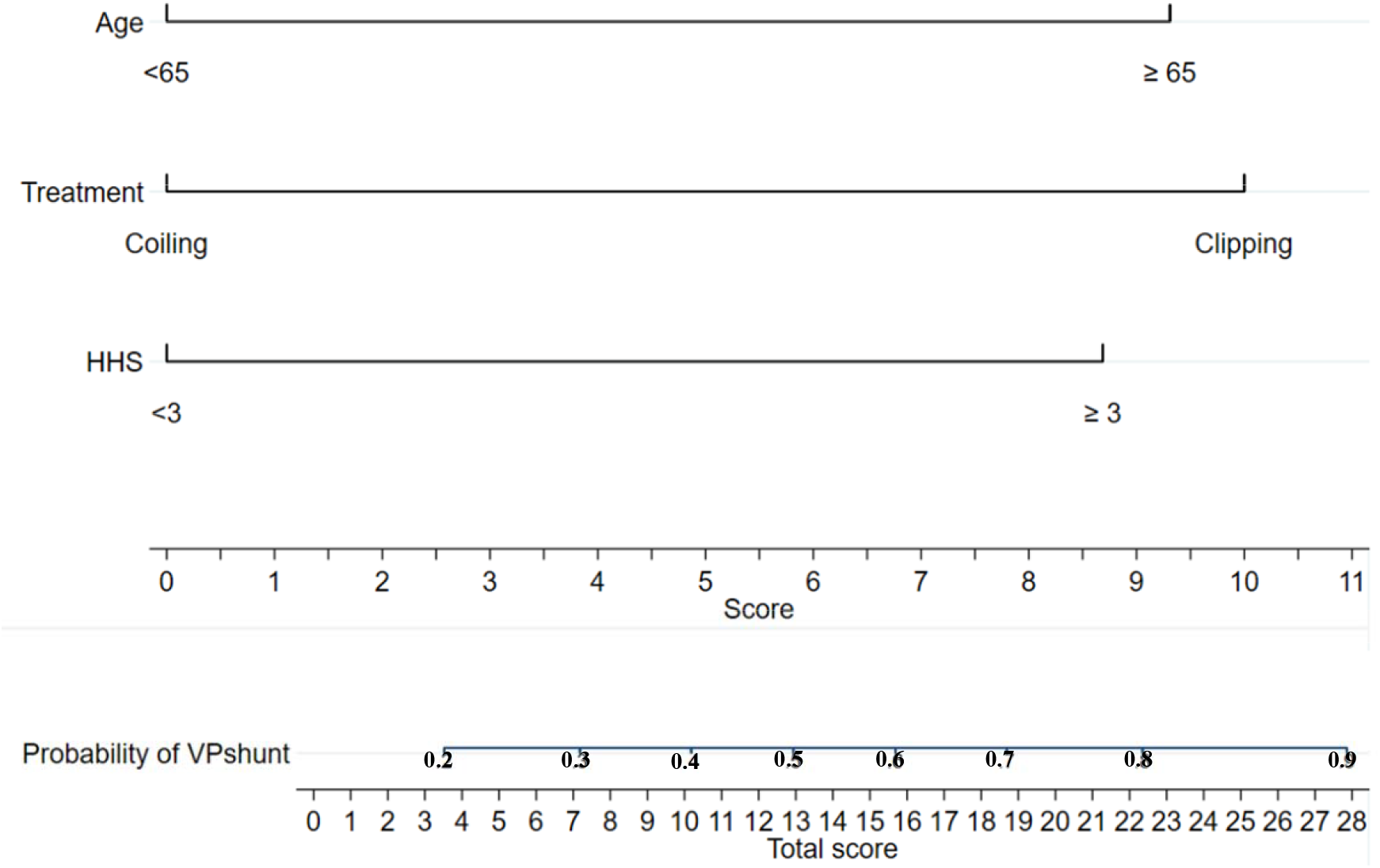
Nomogram plot for predicting SDHC aSAH patients. For an individual patient, each variable corresponds to a point in the fourth row Score: 0-11). The total points can be summed up by points of 3 variables and indicated in the sixth row (the bottom row: 0-28). The probability of SDHC for individual patient can be evaluated by drawing a vertical line from a total point to the fifth row (Probability of VP shunt: 0.2-0.9).

## DISCUSSION

SDHC following aSAH contributes to poor neurological outcome and poses substantial economic impact on health-care system^1^. The incidence of SDHC ranges from 17.2% to 31.2%^3^, correlated factors include old age^4,6,8,14,17,19^, female gender^8^, BMI^10^, high HHS^4,6,8,9,13,15,17,19,28^, high Fisher grade^4,6,7,8,9,15,17,19^, in hospital complication with new neurological deficit^6,8,24,28^, posterior circulation of aneurysms^6,8,28^, acute hydrocephalus^2,6,8,9,13,15,17,19,28^, eGFR^5^ and treatment modality^2,8,14,15,16,17,18,19,20,21,22,23,24,25^.

The SDHC mechanism after aSAH is probably attributed to CSF dynamics alternation^2,3^. Blood products in the subarachnoid space causes arachnoid granulation occlusion and subarachnoid space inflammation, fibrosis and adhesion, which block CSF circulation and absorption^3,5,6,7,8,12,13,14,16,17,18,19^. Additionally, recent studies on animals have demonstrated that CSF production would increase after intraventricular hemorrhage through inflammation-mediated pathway, contributing to the development of post-hemorrhagic hydrocephalus^29,30^.

Several studies have reported that posterior circulation aneurysm was an SDHC risk factor after aSAH, but it was not observed in our cohort study. The supporting evidence databases were mostly before or in the early years of endovascular treatment development^6,8,28^, either all or majority of the patients were treated with clipping surgery in these databases. In our opinion, posterior circulation aneurysms seemed to be significant for SDHC development in patients who underwent clipping surgery rather than endovascular coiling. It is necessary to perform more extensive and more thorough arachnoid dissection to acquire adequate manipulation space in vertebrobasilar aneurysm clipping surgery, which probably led to increased fibrosis and subarachnoid space adhesion and predisposes it to SDHC development after aSAH. In our cohort study, 49% of the cases were treated with endovascular coiling, while only 27% of the cases from the clipping group (15 patients in the clipping group) were posterior circulation aneurysms, this population character may reduce impact of aneurysm location on the development of SDHC.

NLR, PLR, and PNR are reported to be associated with aSAH outcome through the inflammatory and thrombosis activity mechanism^26,27^, but these factors did not contribute to the SDHC development in our cohort.

In contrast to most researches^4,6,7,8,9,15,17,19^, Fisher grade was not shown as an SDHC after aSAH risk factor in our study. We believe it was because every patient in our cohort had EVD placement. The subarachnoid and intraventricular blood was drained out, reduced blood stay within the subarachnoid space, and blood resorption loading. Therefore, this reduced the impact of Fisher grade on SDHC development.

Three risk factors of SDHC after aSAH were identified in our study: age ≥65 years, clipping, and HHS ≥3. Patients with aSAH aged ≥65 years had a higher SDHC rate in comparison with patients aged < 65 years (OR, 3.94; 95% CI, 1.41–11.00; *p* = 0.009). **(Table 3)** A number of studies in regard to SDHC development after aSAH revealed that old age was correlated to SDHC risk^4,6,8,14,17,19^. The result of our study provided more support to this finding that patients age ≥65 years were at significant SDHC risk. Koyanagi et al. assumed that brain atrophy in older patients made the subarachnoid space larger, and can, therefore, harbor more subarachnoid blood burden, which may facilitate inflammation and fibrosis of the subarachnoid space following aSAH^14^. Additionally, we have presumed that a larger subarachnoid space permitted blood to spread more extensively, with resultant inflammation and fibrosis in a more extended area of the subarachnoid space, leading to more obstruction in CSF circulation^3^.

In our study, SDHC frequently occurs in patients with HHS ≥3. Many studies have shown that a high HHS was associated with SDHC^4,6,8,9,10,13,15,17,19,28^. In our study, 59% (31/52) of the patients with high HHS (3–5) underwent a permanent shunt procedure, whereas only 33% (19/57) of patients with low HHS (1-2) was shunt dependent, which showed a strong association between HHS ≥3 and SDHC (OR, 3.59; 95% CI, 1.50–8.61; *p* = 0.004). Our result strengthens the association between high HHS and SDHC after aSAH reported in previous studies.

Our study showed that patients who underwent clipping had a higher SDHC development incidence (OR, 4.36; 95% CI, 1.81–10.53; *p =* 0.001). The treatment modality’s (clipping versus coiling) influence on SDHC remains debated^2,8,14,15,16,17,18,19,20,21,22,23,24,25^. De Oliveira et al. reported that clipping may be associated with a lower risk for developing shunt dependency than coiling in a cohort study that enrolled 596 cases^24^; Loh et al. reported that clipping had a similar ventricular shunt placement incidence compared to coiling in a nationwide database study, which enrolled 10,899 aSAH patients^18^; and Yamada et al. reported that endovascular coiling conferred a lower risk of developing SDHC than microsurgical clipping in a nationwide database study which enrolled 4693 aSAH patients^19^. We found that the study database concluded that clipping was of importance in reducing SDHC risk during the earlier years, especially before 2007. It was noticed that studies enrolled in the database of later years usually reported no difference or coiling superiority on reducing SDHC risk^2,8,14,15,16,17,18,19,21,22,25^. Theoretically, part of subarachnoid clots can be removed during clipping procedures, thus, reduce the CSF circulation obstruction course, which may lead to reduced SDHC risk, but wide dissection of the arachnoid membrane and wide CSF cistern opening during clipping the procedure may increase arachnoid adhesion, which might hinder CSF circulation^22^.

Based on three risk factors and their relative weight identified in our study, we developed a risk nomogram of SDHC development after aSAH **(Figure 1)**, which makes the relative risk of each combination of individual risk factors easily perceptible, which may be helpful in clinical decision making.

The limitations of our study are as follows: 1) it was retrospective and 2) the case number was limited. It is necessary to verify our nomogram in a prospective manner.

## Conclusion

SDHC accounts for 17.2%–31.2% of aSAH patients^3^. The risk factors include age >65 years, ≥3 HHS, and treatment with clipping rather than coiling. A nomogram was developed to make the relative risk of each combination of individual risk factors easily perceptible.

## Data Availability

All data produced in the present work are contained in the manuscript

## Acknowledgments

The authors thank the personnel of the Department of Medical Education and Research of Kaohsiung Veterans General Hospital for providing information, response to inquiries, and assistance in the data processing.

## Contributors

Chia-Ryo Wu wrote the manuscript and Yu-Hone Hsu developed the study. Jin-Shuen Chen, Yao-Shen Chen, and Chun-Hao Yin contributed to the data analysis. Wei-Chuan Liao and Yu-Lun Wu contributed to the data acquisition. Chia-Ryo Wu and Yu-Hone Hsu contributed to the clinical assessment of the data.

## Patient consent for publication

Not required.

## Ethics approval

This study was approved by the Kaohsiung Veterans General Hospital Committee of Human Research and was conducted in accordance with the Declaration of Helsinki 1975. The hospital ethics committee waived the need for patient consent owing to the retrospective observational design and no increase in the health risk of the patients. Grant no: KSVGH22-CT8-03.

## Provenance and peer review

Not commissioned; externally peer reviewed.

## Notes

### Competing Interest Statement

The authors have declared no competing interest.

### Funding Statement

This study did not receive any funding

### Author Declarations

Kaohsiung Veterans General Hospital Committee of Human Research/Grant no: KSVGH22-CT8-03 gave ethical approval for this work

